# Independent serum metabolomics approaches identify disrupted glutamic acid and serine metabolism in Parkinson’s disease patients

**DOI:** 10.1101/2025.06.12.25329524

**Authors:** Jacopo Gervasoni, Carmen Marino, Alberto Imarisio, Lavinia Santucci, Enza Napolitano, Tommaso Nuzzo, Isar Yahyavi, Micol Avenali, Michela Cicchinelli, Gabriele Buongarzone, Caterina Galandra, Marta Picascia, Manuela Grimaldi, Claudio Pacchetti, Francesco Errico, Anna Maria D’Ursi, Andrea Urbani, Enza Maria Valente, Alessandro Usiello

## Abstract

Previous studies assessing blood metabolomic profiles in Parkinson’s disease (PD) patients showed inconsistent results. Here, we employed ¹H-NMR and UPLC/MS analyses on serum samples from a cohort of PD patients and healthy controls (HC). Compared to HC, PD patients showed: (i) higher glutamine, serine, pyruvate and lower α-ketoglutarate levels (^1^H-NMR); (ii) higher glycine and lower glutamic acid concentrations (UPLC/MS). Several pathways associated with amino acids, mitochondrial and antioxidant metabolism emerged as dysregulated in PD. Our findings highlight a prominent disruption of cellular bioenergetic pathways and amino acid homeostasis in PD.

---

Parkinson’s disease (PD) is a common neurodegenerative disorder characterized by highly variable clinical presentation and progression rate. Although recently emerged cerebrospinal fluid (CSF) α- synuclein seed amplification assays have demonstrated high sensitivity and specificity in identifying PD patients^1^, reliable peripheral fluid biomarkers for PD are currently lacking. Beyond nigrostriatal dopaminergic system degeneration, the disruption of cortico-striatal glutamatergic transmission is considered a core feature of PD pathogenesis^2^. We recently evaluated by High-Performance Liquid Chromatography (HPLC) the concentration of a pool of L- and D-amino acids which are known to modulate glutamatergic receptors activation (L-glutamate, L-aspartate, glycine, D-serine) or to represent their precursors (L-glutamine, L-asparagine and L-serine) in the central nervous system (CNS) and blood of PD experimental models and human PD subjects. We demonstrated increased levels of the glutamate N-methyl-D-aspartate receptor (NMDAR) co-agonist D-serine and its precursor L-serine in the (i) rostral putamen of 1-methyl-4-phenyl-1,2,3,6-tetrahydropyridine (MPTP)-treated monkeys^3^, (ii) post-mortem striatum of human PD brains, and (iii) CSF of *de novo* PD patients compared to patients with other neurodegenerative diseases and controls^4^. Finally, we found that the serum D-serine levels positively correlated with age and age at disease onset and were increased in PD patients compared to healthy controls (HC)^5^. Overall, our previous studies showed that PD physiopathology disrupts the central and peripheral homeostasis of serine enantiomers. While these results suggest a putative role for these neuroactive molecules as diagnostic biomarkers in PD, the employed HPLC approach did not allow us to investigate the global metabolic context on which these specific biochemical signatures are grounded. Here, we sought to extend our previous findings by performing untargeted Proton Nuclear Magnetic Resonance (^1^H-NMR)-based metabolomics and targeted ultraperformance liquid chromatography/mass spectrometry (UPLC/MS) analysis on a well-characterized cohort of PD patients whose serum amino acid profile was already reported^5^.

A total of 69 consecutive PD patients and 32 HC were enrolled in the study. The demographic and clinical features of the participants are reported in ***Table 1***. PD and HC were comparable for age, sex distribution, and global cognition. PD group showed a lower Mini Nutritional Assessment (MNA) score, i.e. a slightly higher risk of malnutrition, compared to HC. First, using ^1^H-NMR analysis on the serum of PD and HC groups, we explored the metabolomic profile of the disease. Resonance assignment performed with CHENOMX software detected the presence of 45 metabolites (Supplementary Fig. 1). Metabolite concentrations for each sample were collected in data matrices. Univariate statistical approach was applied using a combined Fold change (FC) and T-test approach. Robust volcano plot analysis evidenced higher serum concentrations of serine, glutamine, creatinine, glycerophosphocholine and pyruvic acid and lower concentrations of 2-oxoglutarate (also named α-ketoglutarate) and acetoacetate in the blood of PD patients compared to HC (Fig. 1a).

**Figure 1.**
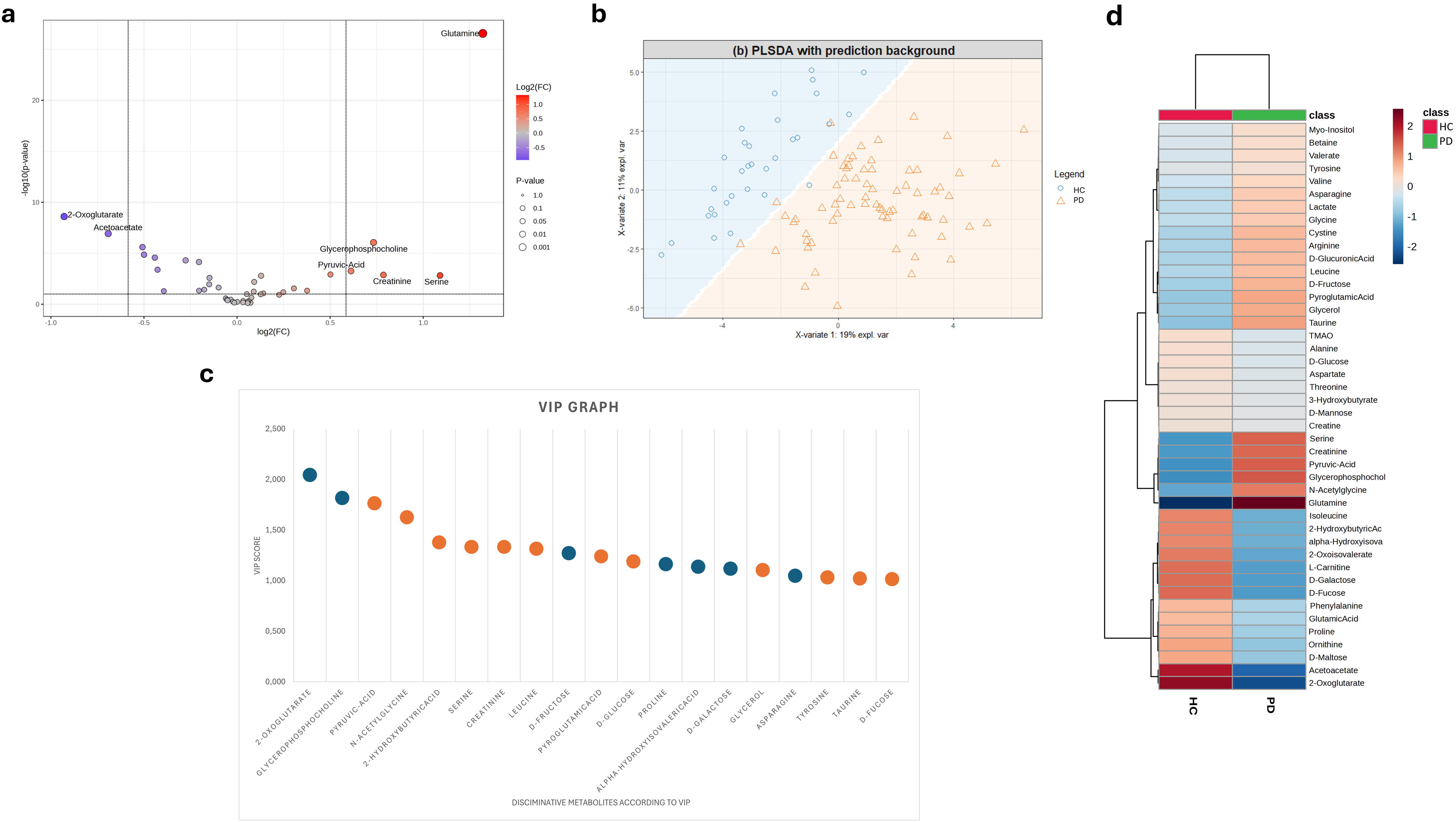
^1^H-NMR metabolomics results. **a** Volcano plot analysis of metabolic changes in PD patients and HC sera. Each point on the volcano plot was based on p-value and fold-change (FC) values, set at 0.05 and 2.0, respectively. Red points identify upregulated metabolites. **b** PLS-DA score scatter plots related to serum from PD patients (N = 69) and healthy controls (N = 32). The cluster analyses are reported in the Cartesian space described by the main components PC1:16.8% and PC2:11.4%. PLS-DA was evaluated using cross-validation (CV) analysis. CV tests performed according to the PLS-DA statistical protocol show a significant cluster separation (0.98 and 0.99 AUC values for PC1 and PC2; see Supplementary Figure 2 for additional details on PLS-DA model performance). **c** VIP score graphs of metabolites discriminating the two clusters. **d.** Heatmap of changed metabolites relative to ^1^H-NMR analyses. The colour of each section corresponds to a concentration value of each metabolite calculated by a normalized concentration matrix (red, upregulated; blue, downregulated). The colour intensity represents the importance of each metabolite in separating the two clusters. Abbreviations: HC, healthy controls; PD, Parkinson’s disease patients.

**Table 1.**
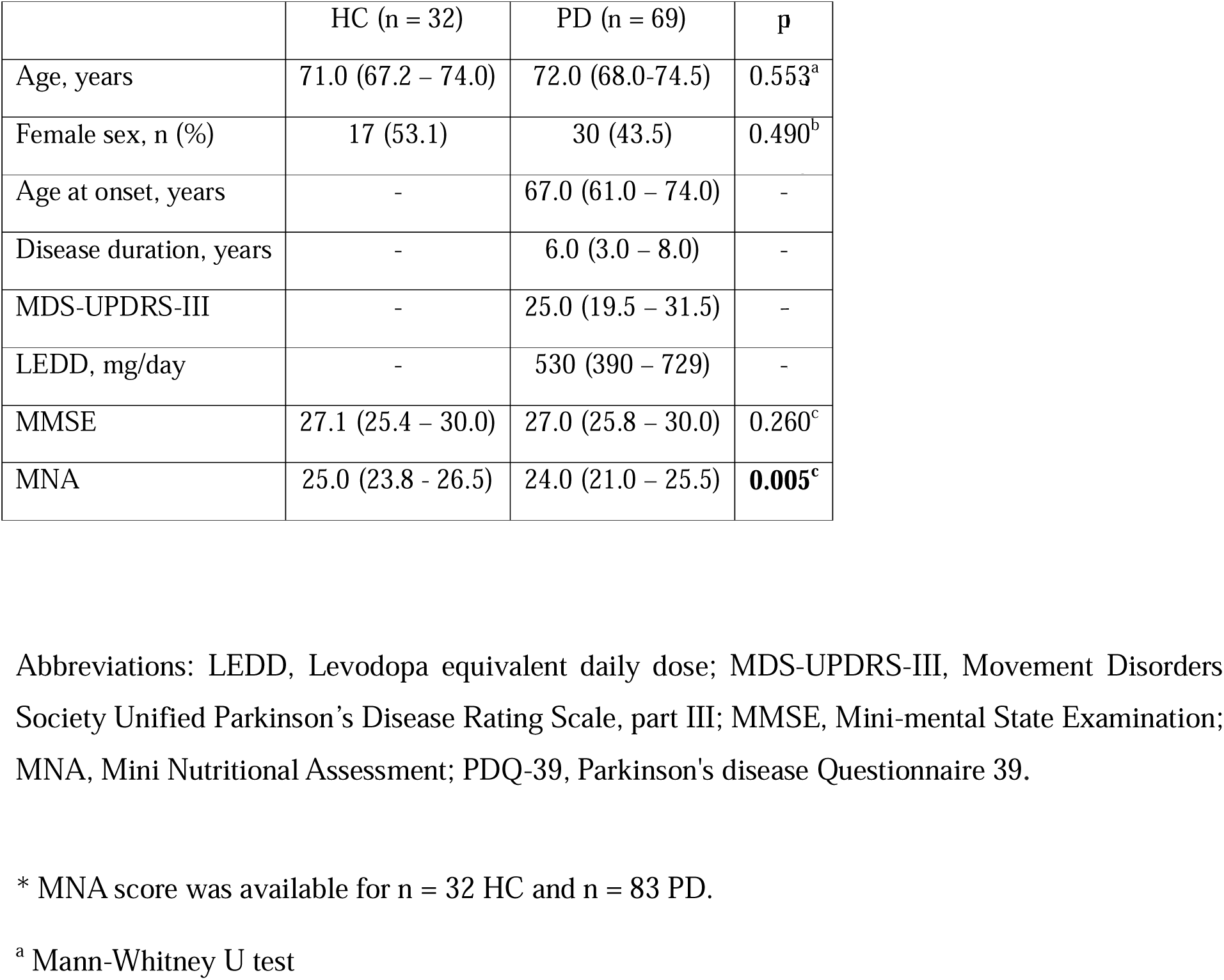
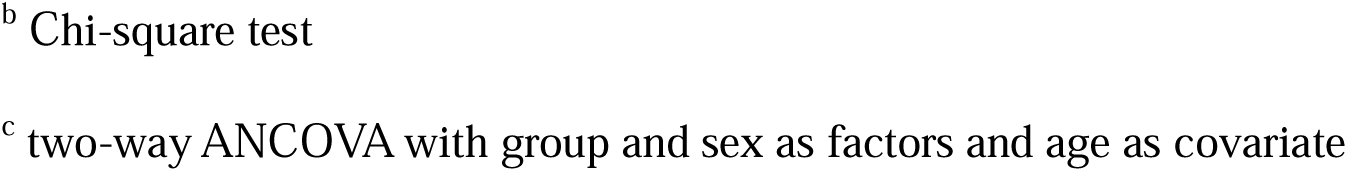
Clinical and demographic features of PD and HC groups enrolled in the study. Data are shown as median (IQR) for continuous variables and sample size (percentage) for categorical variables.

|  | HC (n = 32) | PD (n = 69) | p |
| --- | --- | --- | --- |
| Age, years | 71.0 (67.2 – 74.0) | 72.0 (68.0-74.5) | 0.553<sup>a</sup> |
| Female sex, n (%) | 17 (53.1) | 30 (43.5) | 0.490<sup>b</sup> |
| Age at onset, years | - | 67.0 (61.0 – 74.0) | - |
| Disease duration, years | - | 6.0 (3.0 – 8.0) | - |
| MDS-UPDRS-III | - | 25.0 (19.5 – 31.5) | - |
| LEDD, mg/day | - | 530 (390 – 729) | - |
| MMSE | 27.1 (25.4 – 30.0) | 27.0 (25.8 – 30.0) | 0.260<sup>c</sup> |
| MNA | 25.0 (23.8 - 26.5) | 24.0 (21.0 – 25.5) | **0.005<sup>c</sup>** |

Abbreviations: LEDD, Levodopa equivalent daily dose; MDS-UPDRS-III, Movement Disorders Society Unified Parkinson's Disease Rating Scale, part III; MMSE, Mini-mental State Examination; MNA, Mini Nutritional Assessment; PDQ-39, Parkinson's disease Questionnaire 39.
\* MNA score was available for n = 32 HC and n = 83 PD.
<sup>a</sup> Mann-Whitney U test
<sup>b</sup> Chi-square test
<sup>c</sup> two-way ANCOVA with group and sex as factors and age as covariate

The data matrix was analysed using the multivariate supervised partial least-squares discriminant analysis (PLS-DA) method^6,7^. PLS-DA diagrams indicate that the serum metabolomic profile of PD patients are significantly different from HC (Fig. 1b). The supervised model’s prediction was confirmed by the ROC curve and calculation of error rate (Supplementary Fig. 2)^8,9^. To identify the molecules significantly responsible for metabolomic separation, we performed variable influence on projection (VIP) score analysis^10^. Accordingly, the metabolites characterised by a VIP score > 1 were considered good classifiers between the two clusters^11^. The VIP score graph (***Fig. 1c***) revealed that glycerophosphocholine (VIP: 1.81), N-acetylglycine (VIP: 1.62), creatinine (VIP: 1.33), serine (VIP: 1.33), leucine (VIP:1.31), pyroglutamate (VIP: 1.23), proline (VIP: 1.16), asparagine (VIP:1.04), tyrosine (VIP:1.02) and taurine (1.02) could discriminate the metabolomic profiles of PD patients and HC. Interestingly, VIP analysis showed that the metabolic profiles were also differentiated by several key metabolites related to cellular bioenergetic processes, including 2-oxoglutarate (VIP: 2.04), pyruvic acid (VIP: 1.76), 2-hydroxybutyrate (VIP: 1.37), fructose (VIP: 1.27), glucose (VIP: 1.19) and glycerol (VIP:1.10). Heatmap analysis confirmed the results obtained by robust Volcano plot (Fig. 1d). Reduced concentrations of 2-oxoglutarate and acetoacetate along with an upregulation of creatinine, pyruvic acid, glycerophosphocholine and serine emerged as biochemical blood signatures in PD patients. Additionally, the Receiver Operating Characteristic (ROC) curve confirmed the involvement of 2-oxoglutarate as a mitochondrial-related biomarker in distinguishing PD patients from HC (AUC = 0.94) (Supplementary Fig. 3).

A subsequent enrichment analysis performed on NMR data revealed distinct metabolic pathways dysregulated in PD patients compared to HC, including (i) amino acid pathways, i.e. alanine metabolism, cysteine metabolism, phenylalanine and tyrosine metabolism, glycine and serine metabolism, arginine and proline metabolism, glutamic acid metabolism, aspartate metabolism and tryptophan metabolism; (ii) amino acids catabolism and ammonia recycling, i.e. lysine degradation, valine, leucine and isoleucine degradation, urea cycle, and amino sugar metabolism; (iii) bioenergetic pathways, i.e. glucose-alanine cycle, Warburg effect and malate-aspartate shuttle (Supplementary Fig. 4, Supplementary Table 1).

Since several amino acids emerged among the best discriminating metabolites in ^1^H-NMR analyses, we employed a targeted UPLC/MS approach to better evaluate the serum amino acids profile of PD patients. Of note, UPLC/MS boasts higher sensitivity and resolution compared to ^1^H-NMR^12^. A panel consisting of 44 aminoacidic metabolites was included in UPLC/MS analysis (Supplementary Fig. 5). Supplementary Fig. 6 provides a summary of the common and different metabolites analysed with untargeted ^1^H-NMR and targeted UPLC/MS approaches. Sixteen amino acids were identified in common between the two methodologies.

Univariate analysis on UPLC/MS data between the PD and HC groups highlighted that six amino acids were statistically relevant. Among these, Volcano plot indicates that threonine, glycine, and cystathionine displayed higher levels in PD compared to HC, whereas kynurenine, glutamic acid, and tryptophan were reduced in PD (***Fig 2a***). Based on the univariate analysis results, a PLS–DA classification model between PD and HC was built (***Fig 2b***). A preliminary cross-validation (CV) was performed to determine the optimal number of components. Then, two components were selected for the final PLS-DA model, which was validated using CV. The supervised model’s prediction was confirmed by the ROC curve and calculation of error rate (Supplementary Fig. 7). VIP score analysis, considering even in this case VIP score > 1, identified kynurenine (VIP: 2.17), glutamic acid (VIP: 2.03), tryptophan (VIP: 2.00), cystathionine (VIP: 1.74), threonine (VIP: 1.59), glycine (VIP: 1.47), aspartic acid (VIP: 1.28), arginine (VIP: 1.20), valine (VIP: 1.13), ornithine (VIP: 1.04), lysine (VIP: 1.04), and 4-hydroxyproline (VIP: 1.01) as major contributors in the classification (Fig 2c). Heatmap analysis confirmed the results obtained by Volcano plot (Fig. 2d). An enrichment analysis conducted on UPLC/MS data identified three metabolic pathways that differentiated PD patients from HC: glutathione metabolism, porphyrin metabolism, and tryptophan metabolism (Supplementary Fig. 8).

**Figure 2.**
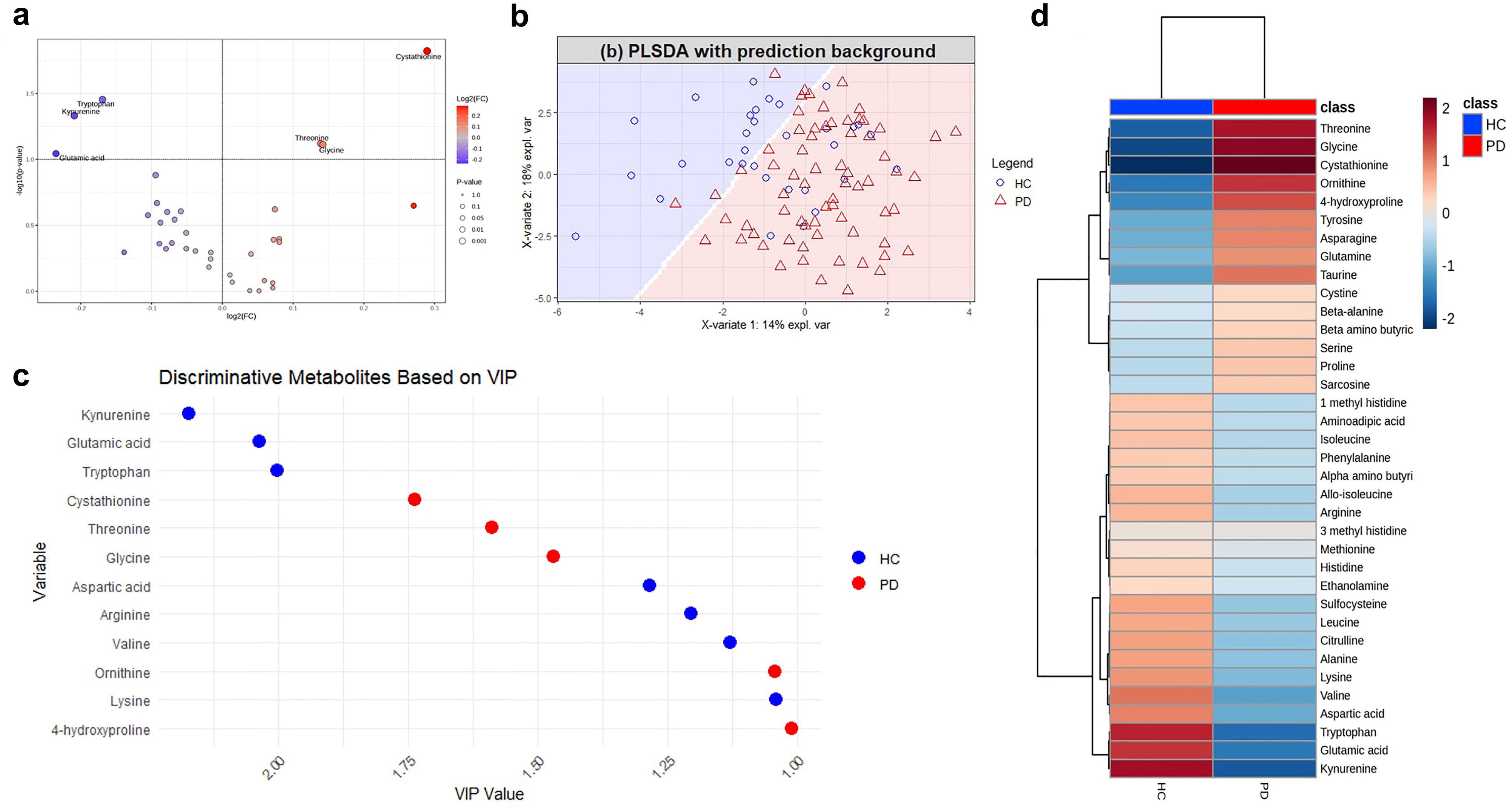
UPLC/MS analysis results **a.** Volcano plot analysis of metabolic changes in PD patients and HC sera. Each point on the volcano plot was based on p-value and fold-change (FC) values, set at 0.1 and 1 respectively. Red points identify upregulated metabolites, and blue points downregulated metabolites. **b** PLS-DA score scatter plots related to serum from PD patients (N = 69) and HC (N = 32). PLS-DA model was evaluated using ROC reporting an AUC value of 0.73 (pvalue: 0.0002) for the first component, and an AUC value of 0.79 (pvalue:3.83 e-6) for the second component. **c.** VIP scores, based on PLS – DA discriminant classification model, were calculated and filtered for a score > 1. Score values are displayed on the x – axis and VIPs on the y – axis. Colors reflect directions of the variables that discriminate the classes. **d.** Heatmap displaying metabolite abundance levels in the PD and HC groups.

Finally, we investigated whether the four metabolites that best discriminated PD from HC correlated with clinical-demographic features in PD patients. In line with our previous HPLC study on PD patients’ serum^5^, we found a positive correlation between glycine levels and motor impairment assessed with the MDS-UPDRS-III score. There were no other significant clinical-biochemical correlations (Supplementary Fig.9).

A recent meta-analysis showed a considerable inconsistency among the findings obtained by 74 original PD-focused metabolomics studies, potentially attributable to the different biospecimens analysed, antiparkinsonian drugs-related effects, different disease stages, genetic background, ethnicity, diet, exercise level, and analytical platform employed^13^. This limited reproducibility highlights the need for deeply phenotyped cohorts to reckon with the several potential confounding factors that may bias the metabolomic profile of PD patients. Moreover, only one prior study used an integrated NMR and MS approach to characterize the plasma metabolomic profile of PD patients^14^. Here, we attempted to untangle this matter adopting a dual approach based on NMR-based metabolomics and UPLC/MS analysis to dissect the serum metabolomic pathways dysregulations in a cohort of PD patients characterized with motor, cognitive, dopaminergic treatment and nutritional assessments. NMR and UPLC/MS analyse different metabolites and employ a different strategy — NMR being untargeted and UPLC/MS concentrating exclusively on amino acids (Supplementary Fig. 6), yet these two methods disclosed variations in metabolites belonging to shared cellular metabolic pathways. In particular, our findings indicate a complex dysregulation of serum amino acids and molecules associated with energy metabolism in PD patients. Consistently, NMR analysis revealed elevated levels of glutamine and serine, accompanied by decreased concentrations of the glutamic acid precursor α-ketoglutarate in PD patients, while UPLC/MS analysis indicated a reduction in serum glutamic acid levels (Figs. 1a, 2a). Notably, both ^1^H-NMR and UPLC/MS revealed altered levels of amino acids closely related to each other. For instance, the lower glutamic acid levels emerged through UPLC/MS in PD patients (Fig. 2a) align well with the reduced α-ketoglutarate concentrations disclosed by NMR analyses (Fig. 1a), overall indicating the occurrence of mitochondrial-related bioenergy abnormalities in PD patients, as reported in previous studies^15^. Another example is represented by the higher serine and glycine concentrations observed in PD group through NMR and UPLC/MS, respectively. Since these two amino acids are interconverted through a single enzymatic reaction catalysed by serine hydroxymethyltransferase^16^, increased serine levels may cause elevated glycine levels and vice versa. Overall, these metabolomic results confirm the homeostasis disruption of the amino acids acting on glutamatergic transmission in the physiopathological framework of PD^2^, and furtherly supports our previous data showing increased serine enantiomers concentration in the striatum of MPTP-lesioned monkeys^3^, post-mortem human PD putamen^4^ and in the CSF^4^ and serum^5^ of PD patients.

The distinct blood metabolomic fingerprint of PD highlighted in this study is also in line with the findings of prior LC/MS^17–21^ and NMR-based^22,23^ studies and supports the idea that PD represents a multi-system clinical-pathological entity rather than a CNS-centred disease. Specifically, the remarkable amino acids dysregulation observed in this study is consistent with previous works reporting altered blood or CSF concentration of glutamic acid, glutamine, glycine and serine in PD patients compared to HC^5,13,22,24,25^. Notably, glutamate is involved in several physiological processes known to be altered in PD, including redox homeostasis, energy metabolism and neuroinflammation^26^. Along with glycine and cysteine, glutamic acid is a key constituent of the antioxidant tripeptide glutathione, and is thus required for maintaining the redox homeostasis. Altered serum glutamate levels may thus ultimately contribute to a dysfunctional glutathione synthesis in PD^17,27,28^. Consistent with this, UPLC/MS data showed a reduction in glutamic acid levels, along with elevated concentrations of glycine, threonine, and cystathionine. This metabolic alteration nicely aligns with the results from UPLC/MS enrichment analysis, which identified glutathione metabolism as one of the most significantly affected pathways (Supplementary Fig. 8), suggesting a disruption in the cellular antioxidant defense in PD. Since glutamic acid is a crucial precursor for glutathione synthesis, we argue that its depletion could impair the capacity to counteract oxidative stress, which is a hallmark of PD pathophysiology^29^.

On the other hand, the increased levels of glycine, cystathionine, and threonine, which are involved in one-carbon and sulfur amino acid metabolism, may reflect a compensatory response to limit oxidative damage or represent a harmful dysregulation in metabolic processes. The positive correlation we observed between serum glycine concentration and MDS-UPDRS-III score supports a putative role for glycine as a peripheral marker of motor dysfunction in PD^5^. Overall, the observed data point to a complex interaction among amino acids, glutathione synthesis, oxidative stress, and the perturbation of key metabolic pathways in PD, highlighting the potential role of these disturbances in disease pathophysiology.

Furthermore, the concomitant downregulation in α-ketoglutarate levels observed in the serum of PD patients suggests a derangement of tricarboxylic acid cycle, which was already reported in other investigations^28,30–32^. This finding, along with (i) the abnormally higher pyruvate concentration; (ii) the perturbation of aerobic glycolysis and glucose-alanine cycle observed in the serum of PD patients, is consistent with the prominent alteration of energy metabolism which characterizes neurodegenerative diseases^15^. In addition to glutathione metabolism, glutamate is pivotal at the intersection of various metabolic pathways critical for energy metabolism, the processing of carbon skeletons (including the citric acid cycle and malate-aspartate shuttle), and ammonia recycling (via the urea cycle). Consistently, our untargeted NMR enrichment analysis showed biochemical alterations in all these energy-related metabolic pathways (Fig. 3 and Supplementary Fig. 4).

**Figure 3.**
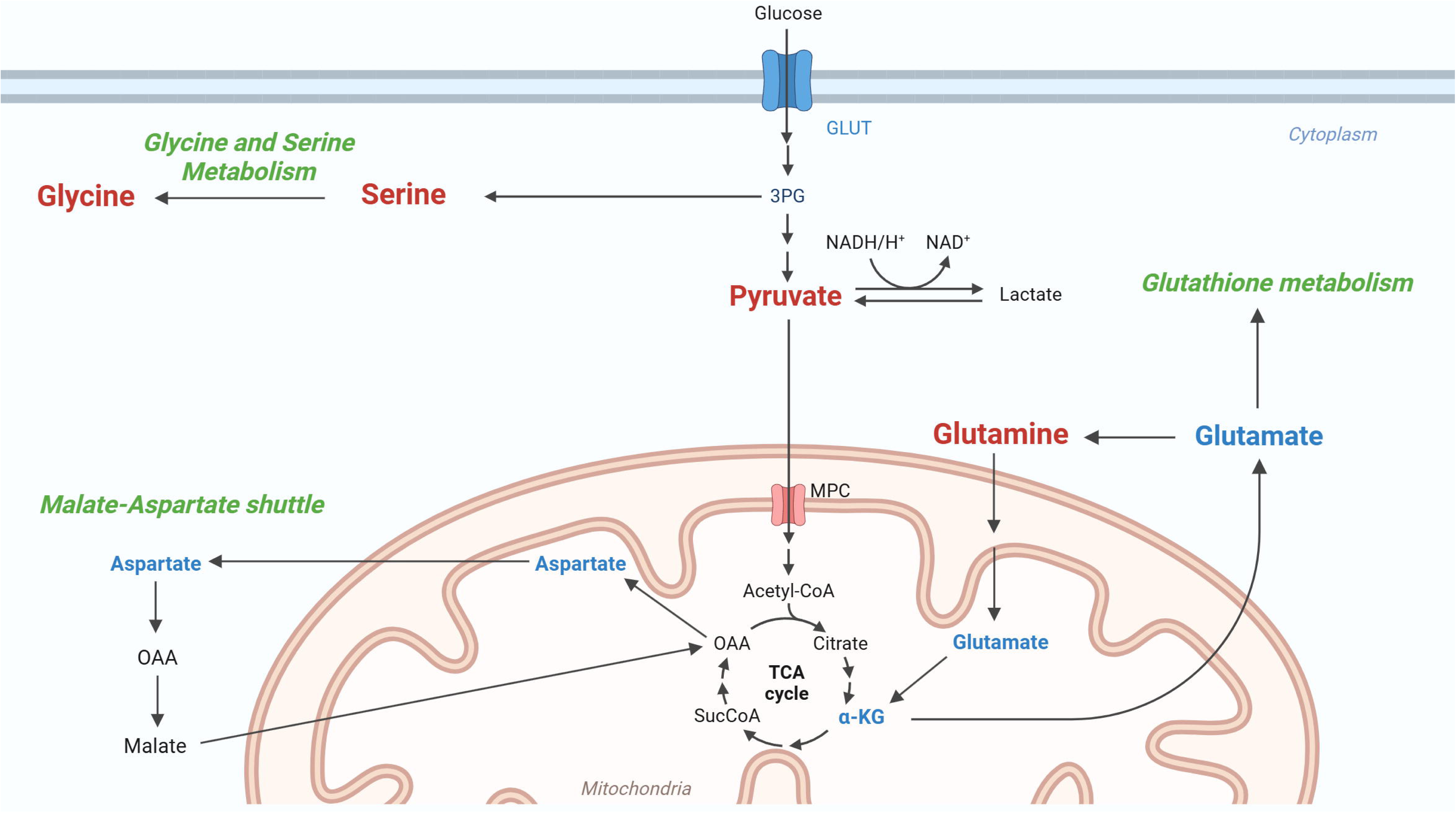
Cartoon depicting the main dysregulated metabolic pathways emerged from ^1^H-NMR and UPLC/MS analyses on PD patients’ serum. Upregulated and downregulated metabolites are shown in red and blue, respectively. Metabolic pathways over-represented in PD are labelled in green. Abbreviations: Acetyl-CoA, acetyl coenzyme A; α-KG, α-Ketoglutarate; GLUT, glucose transporter; MCT, Monocarboxylate transporter; MPC, Mitochondrial pyruvate carrier; OAA, Oxaloacetate; 3PG, 3-Phosphoglyceric acid; SucCoa, Succinyl-coenzyme A; TCA, Tricarboxylic Acid Cycle. Created with Biorender.com.

Finally, in PD patients we observed a decrease in both kynurenine and tryptophan levels, key metabolites in the tryptophan-kynurenine pathway, which is crucial for neuroprotection and immune modulation^33^. This reduction, combined with enrichment analysis revealing alterations in tryptophan metabolism, is consistent with growing experimental evidence showing a significant dysregulation of the tryptophan-kynurenine pathway in PD^34,35^. Kynurenine is involved in the biosynthesis of neuroprotective metabolites like kynurenic acid, which modulates glutamate activity and inflammation^33^. The concomitant depletion of tryptophan and kynurenine may indicate impaired neuroprotective mechanisms or increased oxidative stress, aligning with the altered tryptophan metabolism observed with both NMR and UPLC/MS (Supplementary Figs. 4, 8).

Dietary regimen may also affect the blood metabolomic profile^13^. According to the MNA assessment, 34 (40.4%) of the PD patients enrolled in this study were at risk for malnutrition (i.e., MNA score < 23.5) versus only 5 (14.7%) HC. Although the impact of malnutrition on peripheral metabolome has been poorly characterized in PD^36^, differences in diet between the two groups may have partially contributed to the metabolomic signatures observed in our study. However, the lack of correlation between MNA score and the serum levels of the most discriminative metabolites identified through multivariate UPLC/MS analyses (Supplementary Fig. 9a), as well as the overnight fasting preceding serum sampling, support a main contribution of PD physiopathology in determining the metabolic alterations observed in this study.

The primary limitation of the current cross-sectional study is the unequal number of controls compared to PD patients.

In conclusion, our study highlights disrupted homeostasis of molecules related to glutamic acid, serine and energy metabolism as distinct serum signatures in PD patients. Analysis of the serum metabolome in populations at high risk of conversion to PD, such as subjects with idiopathic REM sleep behavior disorder or asymptomatic carriers of genetic risk variants, is warranted to assess its value as an early diagnostic biomarker.

## Methods

### Participants

Eighty-eight consecutive patients with a clinical diagnosis of PD^37^ with disease duration of at least one year and sustained dopaminergic treatment response were consecutively recruited at IRCCS Mondino Foundation, Pavia, Italy, between January 2019 and December 2021. Thirty-four HC were selected among patients’ caregivers and were subjected to a complete neurological examination and cognitive screening (Mini-Mental State Examination, MMSE) to exclude parkinsonian sign and cognitive impairment, respectively. This study was approved by the local ethics committee (protocol 20180097520, 09/11/2018) and was in conformity with the Helsinki Declaration. Written informed consent was obtained from all participants. All procedures were performed in compliance with relevant laws and institutional guidelines.

All PD patients underwent (i) brain magnetic resonance imaging in order to exclude prominent cortical/subcortical infarcts, cerebral small vessel disease or atypical signs (such as midbrain, cortical or cerebellar atrophy) indicating atypical parkinsonism; (ii) ^123^I-FP-CIT SPECT imaging to confirm nigrostriatal dopaminergic degeneration. Each patient underwent a standardized neurological examination, including the motor assessment with the Movement Disorders Society Unified Parkinson Disease Rating Scale part III (MDS-UPDRS-III), global cognition (MMSE) and Mini Nutritional Assessment^38^. Levo-dopa equivalent daily dose (LEDD) was also calculated at baseline according to the last proposed conversion factors^39^. Patients presenting with dementia according to current criteria^40^ were excluded from the study. The following exclusion criteria were also applied: (1) Hoehn and Yahr stage > 3; (2) diagnosis of atypical parkinsonism including corticobasal syndrome (CBS), progressive supranuclear palsy (PSP), multiple system atrophy (MSA), dementia with Lewy bodies (DLB); (3) any systemic condition potentially affecting serum amino acid levels, including kidney, liver, rheumatologic and neoplastic diseases, history of drug or alcohol abuse; (4) history of altered serum creatinine levels (> 1.2 mg/dl) or liver function parameters (aspartate transaminase or alanine transaminase > 50 U/l). To ensure the absence of potential confounding factors, exclusion criteria no. 3 and 4 were also applied to HC cohort.

### Samples preparation

Blood sampling was performed after an overnight 12-h fasting and antiparkinsonian treatment washout period. Serum was collected according to Standard Operating Procedure (SOP)^41^ to perform NMR-based metabolomic analysis. In order to remove excess proteins, the serum was filtered using Amicon Ultra-0.5 3000 MWCO pre-rinsed (washed 7 times) at 4 °C using a centrifuge (force 12,000 g). Before NMR spectroscopy measurements, the blood serum was aliquoted and stored at −80°C in Greiner cryogenic vials^41^. NMR samples were prepared by adding 250 μL of phosphate buffer to 250 μL of filtered sera, including 0.075 M Na_2_HPO_4_ x7 H_2_O, 4% NaN_3_, and H_2_O. Trimethylsilyl propionic-2,2,3,3-d_4_ acid, sodium salt (TSP 0.1% in D_2_O) was used as an internal reference for the alignment and quantification of NMR signals; the mixture, homogenized by vortexing for 30 sec, was transferred to a 5 mm NMR tube (Bruker NMR tubes) before acquisition^41^.

### NMR data acquisition, processing and assignment

NMR experiments were acquired for all samples on a Bruker Ascend™ 600 MHz spectrometer equipped with a 5 mm triple resonance Z gradient TXI probe (Bruker Co, Rheinstetten, Germany) at 298 K. TopSpin, version 3.2 was used for the spectrometer control and data processing (Bruker Biospin). CPMG (Carr□Purcell–Meiboom–Gill) experiments were performed on serum samples and acquired using 20 ppm spectral width, 32 k data points, with f1 presaturation and T2 filter using D20 of 300 µsec, D1 of 4 sec. A weighted Fourier transform was applied to the time domain data with a line widening of 0.5 Hz followed by a manual step and baseline correction in preparation for targeted profiling analysis. Assignment of ^1^H-NMR signals performed with Chenomix software^42^ on 1D ^1^H CPMG NMR spectra detected the presence of 45 metabolites (Supplementary Fig. 1). The quantification (concentrations in µM) was carried out using automated ASICS^43^.

### UPLC/MS methods

The concentrations of a panel of 44 amino acids and derivatives were measured on serum samples by UPLC/MS. The panel includes: 1-Methylhistidine, 3-Methylhistidine, 4-Hydroxyproline, α- Aminobutyric acid, β-Alanine, β-Aminobutyric acid, γ-Aminobutyric acid, Alanine, Allo-Isoleucine, Aminoadipic acid, Anserine, Arginine, Asparagine, Aspartic acid, Carnosine, Citrulline, Cystathionine, Cystine, Ethanolamine, Glutamic acid, Glutamine, Glycine, Glycil proline, Histidine, Homocitrulline, Homocysteine, Hydroxylysine, Isoleucine, Kynurenine, Leucine, Lysine, Methionine, Ornithine, Phenylalanine, Phosphoethanolamine, Proline, Sarcosine, Serine, Sulfocysteine, Taurine, Threonine, Tryptophan, Tyrosine, Valine. Briefly, 50 µL of the sample were added to 100 µL 10% (w/v) sulfosalicylic acid containing an internal standard mix (50 µM) (Cambridge Isotope Laboratories, Inc., Tewksbury, MA, USA). The mixture was centrifuged at 10000 rpm for 15 min. 70 µL of borate buffer and 20 µL of AccQ Tag reagents (Waters Corporation, Milford, MA, USA) were added to 10 µL of the obtained supernatant and heated at 55 °C for 10 min. Next, samples were loaded onto a CORTECS UPLC C18 column 1.6 µm, 2.1 mm x 150 mm (Waters Corporation) for chromatographic separation (ACQUITY H-Class, Waters Corporation). Elution was accomplished at 0,5 mL/min flow-rate with a linear gradient (9 min) from 99:1 to 1:99 water 0.1% formic acid/acetonitrile 0.1% formic acid. Analytes were detected on an ACQUITY QDa single quadrupole mass spectrometer equipped with an electrospray source operating in positive mode (Waters Corporation).

The analytical process was monitored using amino acid controls (level 1 and level 2) manufactured by the MCA laboratory of the Queen Beatrix Hospital (The Netherlands). Serum amino acid concentrations were determined by comparison with values obtained from a standard curve for each amino acid (2,5-10-50-125-250-500 μmol/L only for Cysteine, 5-20-100-250-500-1000 μmol/L for all amino acids). For data analysis (calibration curves and amino acid quantitation), the instrument software TargetLynx was used.

### Clinical-demographics statistical analysis

Clinical and demographic characteristics were described using, as summary statistics, median and the interquartile range (IQR) or absolute and relative frequencies. Comparisons between PD patients and HC were evaluated using Mann Whitney U test for continuous variables and Chi-Square test for dichotomous variables. Between-group comparisons of MMSE and MNA scores were evaluated with two-way independent ANCOVA with “group” and “sex” as factors and “age” as covariate.

### ^1^H-NMR and UPLC/MS statistical analysis

Before statistical and bioinformatic analyses, data obtained from mass spectrometry were processed to construct the final dataset. The following condition was applied for variable (amino acid) inclusion: variables that were not detected or had more than 50% missing values were excluded. Based on this threshold, 36 variables were selected, and the missing values were imputed using the PPCA method in Metaboanalyst 6.0.

The data matrices were normalised prior to the application of the biostatistical method. Specifically, a logarithmic transformation was applied to the NMR data matrix, and an autoscaling normalization was applied to UPLC/MS data. A univariate analysis was performed on both the LC-MS and NMR data matrices utilising a Robust Volcano plot determined using the FC, calculated as the ratio of PD/HC, with a threshold set at FC=1. Volcano plots were made using the FC= 1 and p-value < 0.1 as thresholds using MetaboAnalyst 6.0.

Multivariate classification model based on PLS–DA was built on both UPLC/MS and NMR data to define similarities and differences between the PD and HC groups. PLS-DA analysis was conducted using the mixOmics (6.30.0) R package. VIP scores > 1 were represented as dot plots generated using ggplot2, dplyr, and tidyr. The best number of components was calculated using a 10-fold cross-validation to minimize model’s error. Model performance was validated using a 10-fold cross-validation with 50 iterations. VIP scores were calculated to assess the importance of each variable to class separation.

The validation of the supervised models involved calculating the area under the curve and assessing the error rate through Balanced Error Rate (BER) and overall error (OVERALL) metrics computed on the first and second components using maximum, centroid, and Mahalanobis distance^44^. To provide an intuitive view of the data, we performed heatmaps using normalized data, average group concentration, Euclidian distance and Ward method^45^. Biomarker analysis was carried out by analysing the univariate ROC curve to calculate AUC and its 95% confidence intervals (500 bootstrap cycles methods)^46^. The analysis of the Kyoto Encyclopedia of Genes and Genomes (KEGG) pathways was carried out using Enrichment tool of MetaboAnalyst. Biochemical pathways with FDR-adjusted p-values below 0.05 and a hit value (i.e., the number of metabolites in the pathways) exceeding 1 were taken into account.

The correlation between the UPLC/MS metabolites showing VIP score > 1.0 and PD patients’ clinical-demographic features was plotted as a correlation matrix showing Pearson’s correlation coefficients and p-values obtained using the MATLAB Statistics and Machine Learning Toolbox (R2024a version, MathWorks Inc., Natick, MA, USA) and MATLAB corrcoef function (R2024b). Data were previously subjected to average normalization. Scatterplots of significative correlations were executed using MATLAB. Original and normalized (log – transformation) abundances were displayed.

## Data availability

Metabolomics data have been deposited to the EMBL-EBI MetaboLights database (https://www.ebi.ac.uk/metabolights/) with the identifier MTBLS10958. UPLC/MS data have been deposited to the MassIVE database (https://massive.ucsd.edu/) with the identifier MSV000097026.

## Acknowledgments

This study was partially funded by CARIPLO Foundation (grant nr. 2017-0575 to EMV and AU), Italian Ministry of Health (Ricerca Corrente 2022-2024 to IRCCS Mondino Foundation) and Italian Ministry of University and Research (PRIN 2022 - COD. 2022XF7YYL_02 to AU). The work of E.M.V. and A.U. is supported by #NEXTGENERATIONEU (NGEU) and funded by the Ministry of University and Research (MUR), National Recovery and Resilience Plan (NRRP), project MNESYS (PE0000006) – A Multiscale integrated approach to the study of the nervous system in health and disease (DN. 1553 11.10.2022).

The authors are grateful to all the patients, their caregivers and the Clinical Trial Center of IRCCS Mondino Foundation for the kind cooperation with this study.

## Author contributions

**JG:** Data curation; Formal analysis; Writing - original draft; Writing - review & editing. **CM**: Data curation; Formal analysis; Writing - original draft; Writing - review & editing. **AI**: Data curation; Writing - original draft; Writing - review & editing; **LS:** Data curation; Formal analysis. **EN**: Formal analysis. **TN**: Data curation. **IY**: Writing - review & editing. **MA**: Writing - review & editing. **MC:** Data curation; Formal analysis; **CG**: Investigation. **GB**: Investigation. **CG**: Investigation. **MP**: Investigation. **MG**: Data curation; Formal analysis. **CP**: Writing - review & editing. **FE**: Writing - review & editing. **AMD**: Project administration; Supervision; Writing - review & editing. **AUr:** Project administration; Supervision. **EMV**: Conceptualization; Funding acquisition; Project administration; Resources; Supervision; Writing - review & editing. **AUs**: Conceptualization; Funding acquisition; Project administration; Resources; Supervision; Writing - review & editing. All authors read and approved the final manuscript.

