## Supplementary material for "Independent serum metabolomics approaches identify disrupted glutamic acid and serine metabolism in Parkinson’s disease patients"

**Supplementary Table 1** Quantitative enrichment analysis performed on $^1$H-NMR and related to PD and HC serum metabolomic results. Hits represent the number of metabolites involved in the pathways and specified in the 'Metabolites' column. The pathways were considered statistically significant with Hits >1, p-value < 0.05, adjusted p-value performed using the Holm-Bonferroni test (Holm p) and False Discovery Rate (FDR) < 1.

| | hit | Raw p | Holm p | FDR | Metabolites |
|---|---|---|---|---|---|
| Alanine Metabolism | 5 | 3,10E-16 | 2,05E-14 | 1,64E-14 | Glutamate, Glutamine, Pyruvate, Oxoglutarate, Alanine |
| Amino Sugar Metabolism | 5 | 4,97E-16 | 3,23E-14 | 1,64E-14 | Glutamate, Pyruvate, Glutamine, Fructose, Glutamine |
| Propanoate Metabolism | 4 | 9,39E-16 | 6,01E-14 | 1,77E-14 | Glutamate, Oxoglutarate, 2-Hydroxybutyrate, Valine |
| Cysteine Metabolism | 3 | 1,07E-15 | 6,77E-14 | 1,77E-14 | Glutamate,Glutamine, Pyruvate, Oxoglutarate |
| Glucose-Alanine Cycle | 6 | 2,26E-15 | 1,40E-13 | 2,89E-14 | Glutamate, Glutamine, Pyruvate, Oxoglutarate, Alanine,Glucose |
| Warburg Effect | 7 | 1,82E-14 | 1,05E-12 | 1,33E-13 | Glutamate, Pyruvate, Oxoglutarate, Glucose, Lactate, Glutamine |
| Phenylalanine and Tyrosine Metabolism | 5 | 4,16E-14 | 2,37E-12 | 2,75E-13 | Glutamate, Oxoglutarate, Acetoacetate, Phenylalanine,Tyrosine |
| Tyrosine Metabolism | 5 | 7,66E-14 | 4,29E-13 | 4,60E-13 | Glutamine, Oxoglutarato,Acetoacetato,Aspartate, Tyrosine |
| Glycine and Serine Metabolism | 10 | 1,66E-13 | 9,15E-12 | 8,72E-13 | Glutamate, Pyruvate, Oxoglutarate, Serine, Ornithine, Betaine, Alanine,Threonine, Arginine, Creatine |
| Arginine and Proline Metabolism | 7 | 1,80E-13 | 9,72E-12 | 8,72E-13 | Glutamate, Proline,Oxoglutarate,Ornithine,Aspartate,Arginine,Creatine |
| Ammonia Recycling | 8 | 1,85E-13 | 9,80E-12 | 8,72E-13 | Glutamate,Pyruvate,Oxoglutarate,Aspartate,Glutamine, Alanine,Glutamine |
| Glutamate Metabolism | 7 | 2,48E-13 | 1.29e-15 | 1,09E-12 | Glutamate,Pyruvate,Oxoglutarate,Serine,Aspartate,Glutamine,Asparagine |
| Beta-Alanine Metabolism | 3 | 2,92E-13 | 1,49E-11 | 1,14E-12 | Glutamate,Oxoglutarate,Aspartate |
| Malate-Aspartate Shuttle | 3 | 2,92E-13 | 1,49E-11 | 1,14E-12 | Glutamate,Oxoglutarate,Aspartate |
| Lysine Degradation | 2 | 3,54E-13 | 1,74E-11 | 1,30E-12 | Glutamate,Oxoglutarate |
| Tryptophan Metabolism | 3 | 4,88E-13 | 2,34E-11 | 1,69E-12 | Glutamate,Oxoglutarate, Alanine |
| Urea Cycle | 8 | 8,59E-13 | 4,04E-11 | 2,84E-13 | Glutamate, Pyruvate,Oxoglutarate, Alanine,Arginine,Aspartate,Ornithine,Glutamine |
| Purine Metabolism | 3 | 9,70E-12 | 4,46E-10 | 3,05E-11 | Glutamate,Aspartate,Glutamine |
| Valine, Leucine and Isoleucine Degradation | 7 | 2,66E-11 | 1,17E-09 | 7,64E-11 | Glutamate, Ketosisovalerate, Oxoglutarate, Isoleucine,Leucine,Acetoacetate,Valine |
| Aspartate Metabolism | 6 | 1,05E-09 | 4,50E-08 | 2,88E-09 | Glutamate,Aspartate,Glutamine,Asparagine,Arginine, 2-Oxoglutarate |

**Supplementary Table 2.** Pathway enrichment analysis performed on UPLC/MS data and related to PD and HC serum metabolomic results. Hits represents the numbers of metabolites involved in the pathways and specified in the 'Metabolites' column. The pathways were considered statistically significant with Hits >1, p-value < 0.05, adjusted p-value performed using Holm Bonferroni test (Holm p) and False Discovery Rate (FDR) < 1.

| | hit | Raw p | Holm p | FDR | Metabolites |
|---|---|---|---|---|---|
| Tryptophan metabolism | 2 | 1.12E-02 | 3.71E-01 | 3.71E-01 | Tryptophan, Kynurenine |
| Porphyrin metabolism | 2 | 3.33E-02 | 1.00E+00 | 3.77E-01 | Glycine, Glutamic Acid |
| Glutathione metabolism | 3 | 3.43E-02 | 1.00E+00 | 3.77E-01 | Glycine, Glutamic Acid Ornithine |

**Supplementary Figure 1.** Representative 1D [1] H CPMG spectrum of PD patient serum. The spectrum was acquired at 600 MHz and dT = 298 K. Fourty-four metabolites were identified and annotated as follows: 1: 2-Hydroxybutyrate; 2: 2-Hydroxyisovalerate; 3: 2-Oxoglutarate; 4: 2- Oxoisovalerate; 5: 3-Hydroxybutyrate; 6: Acetoacetate; 7: Alanine ; 8: Arginine; 9: Asparagine; 10:Aspartate; 11: Betaine; 12: Carnitine; 13: Creatine; 14: Creatinine; 15: Cystine; 16: Fructose; 17: Fucose; 18: Galactose; 19: Glucose; 20: Glucuronate; 21: L-Glutamic acid 22; L-Glutamine; 23:Glycerol; 24: Glycine; 25: Lactate; 26: Leucine; 27: Isoleucine; 28: Maltose; 29: Mannose; 30: Myo-inositol; 31: N-acetylGlycine; 32: Ornithine; 33: Phenylalanine; 34: Proline; 35: Pyroglutamate; 36: Pyruvate; 37:Serine; 38: Glycero-3-phophocholine; 39: Taurine; 40: Threonine;41: TMAO; 42: Tyrosine; 43: Valeric acid; 44: Valine; 45:Methanol.

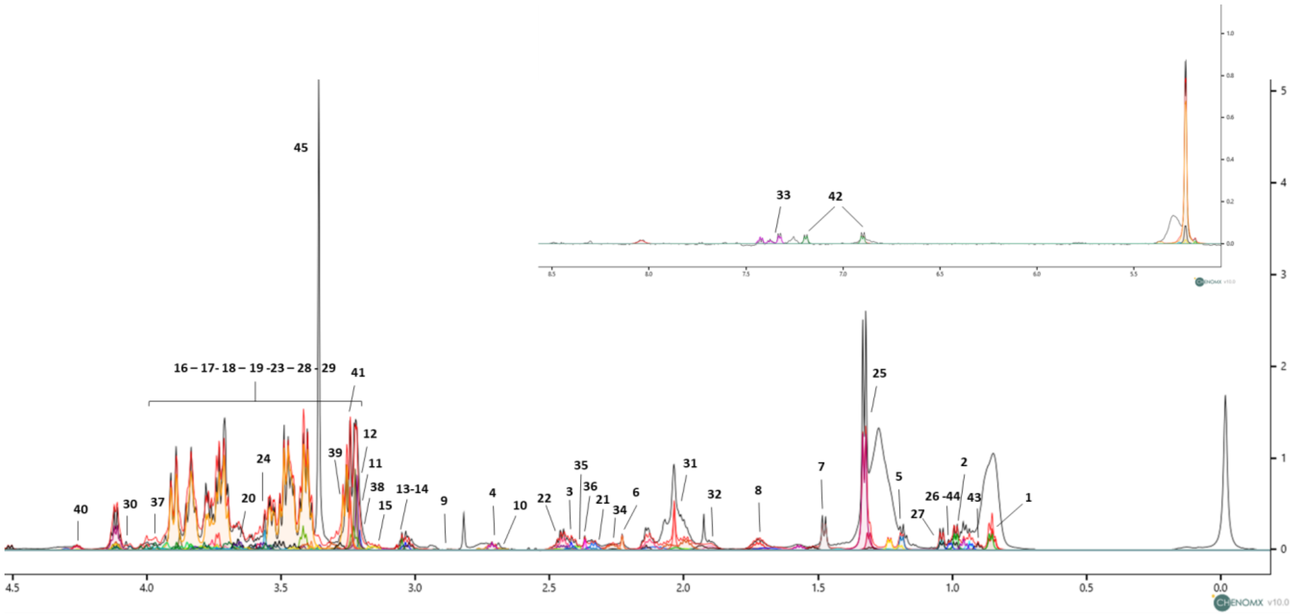


**Figure: 1:** 2-Hydroxybutyrate; **2:** 2-Hydroxyisovalerate; **3:** 2-Oxoglutarate; **4:** 2-oxoisovalerate; **5**: 3-Hydroxybutyrate; **6:** Acetoacetate; **7:** Alanine ; **8:** Arginine; **9:** Asparagine; **10:** Aspartate; **11:** Betaine; **12:** Carnitine; **13:** Creatine; **14:** Creatinine; **15:** Cystine; **16:** Fructose; **17:** Fucose; **18:** Galactose; **19:** Glucose; **20:** Glucuronate; **21:** L-Glutamic acid; **22:** Glutamine; **23:**Glycerol; **24:** Glycine; **25:** Lactate; **26:** Leucine; **27:** Isoleucine; **28:** Maltose; **29:** Mannose; **30:** Myo-inositol; **31:** N-acetylGlycine; **32:** Ornithine; **33:** Phenylalanine; **34**: Proline; **35**: Pyroglutamate; **36**: Pyruvate; **37**:Serine; **38**: Glycero-3-phophocholine; **39**: Taurine; **40**: Threonine; **41**: TMAO; **42**: Tyrosine; **43**: Valeric acid; **44**: Valine; **45**:Methanol

**Supplementary Figure 2. a**. Performance evaluations of the PLS-DA model related to $^{1}$H-NMR data carried out using ROC curve. The ROC curve values for the first and second components with the relative p-value are given in Table **b**. Balanced error rate (BER) and total error (OVERALL) are calculated on the first and second components using maximum, centroid and Mahalanobis distance.

**a**

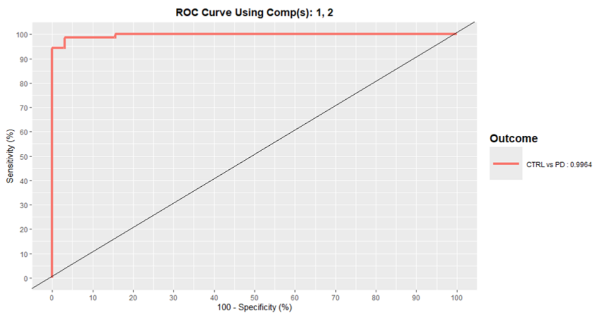


| AUC Component 1 | p-value |
|---|---|
| 0.98 | 1.34e-12 |
| **AUC Component 2** | **p-value** |
| 0.99 | 1.11e-15 |

**b**

| BER | MAX.DIST | CENTROID .DIST | MAHALANOBIS. DIST |
|---|---|---|---|
| COMP1 | 0,18 | 0,17 | 0,17 |
| COMP2 | 0,05 | 0,09 | 0,07 |
| **OVERALL** | **MAX.DIST** | **CENTROID .DIST** | **MAHALANOBIS. DIST** |
| COMP1 | 0,14 | 0,17 | 0,17 |
| COMP2 | 0,06 | 0,10 | 0,09 |

**Supplementary Figure 3.** Receiver operating characteristic (ROC) curves related to 2-oxoglutarate serum concentration. The Cartesian space is described by x-axis: false positive rate and y-axis: true positive rate. ROC curve has two components, the empirical ROC curve that is obtained by joining the points represented by the sensitivity and the specificity for the different cutpoints and the chance diagonal represented by the 45-degree line drawn through the coordinates (0,0) and (1,1).

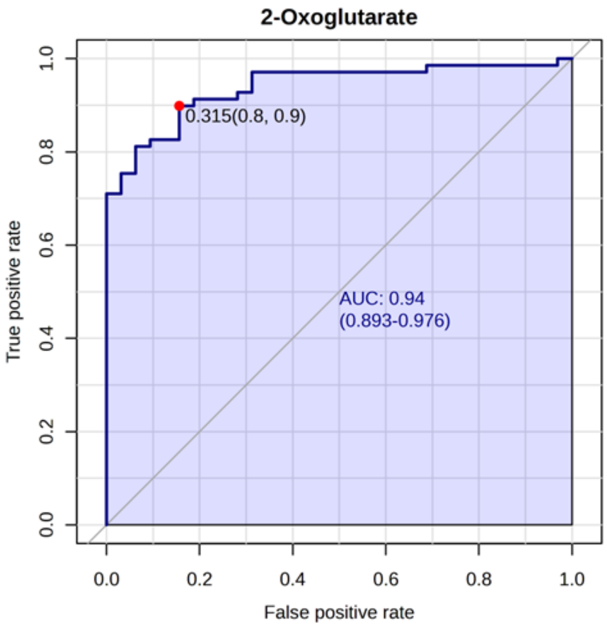

**Supplementary Figure 4.** Enrichment pathways analysis performed on PD and HC metabolites identified by $^{1}$H-NMR using Metaboanalyst 6.0; the discriminative pathways are ranked according to p-value and number of hits reported in the bars.

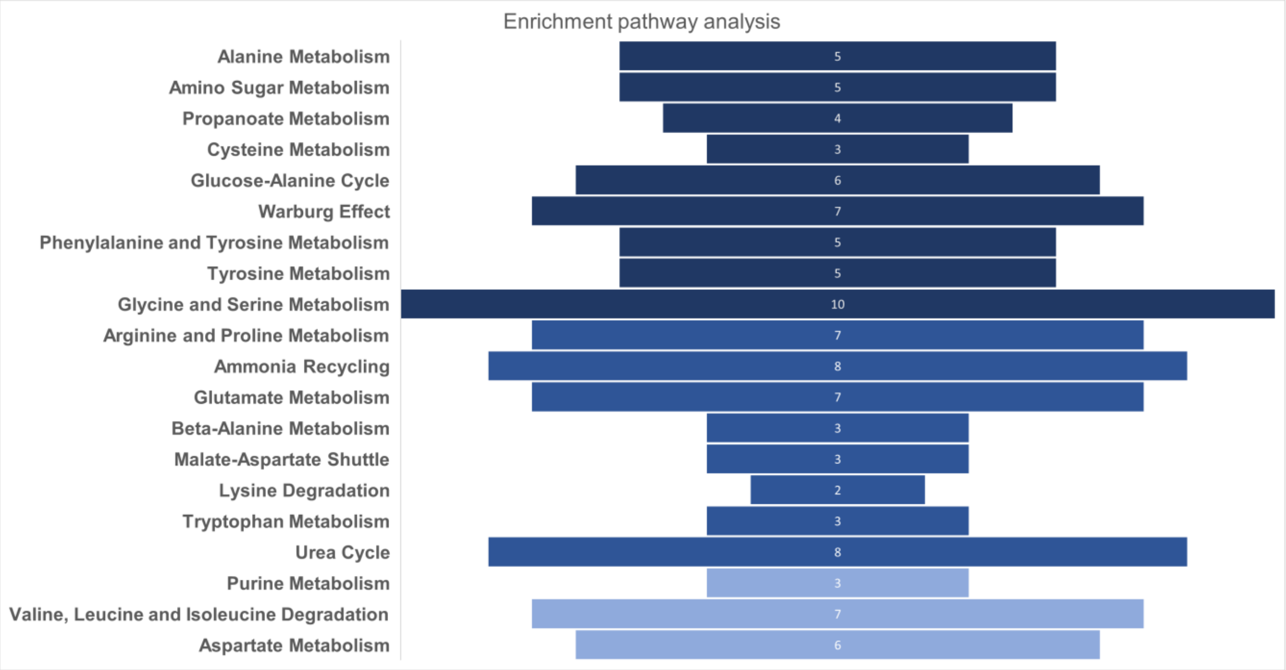

**Supplementary Figure 5.** Representative MS Total Ion Current (TIC) of PD patients' serum. The spectrum was acquired in Selected Ion Recording (SIR), and forty-four amino acids were quantified. Peak annotation as follows: **1:** histidine and histidine-IS; **2:** 3-methyl-histidine; **3:** hydroxyproline; **4:** 1-methyl-histidine; **5:** asparagine and asparagine-IS; **6:** arginine and arginine-IS; **7:** carnosine; **8:** phosphoethanolamine; **9:** anserine; **10:** taurine; **11**:serine and serine-IS; **12:** glutamine and glutamine-IS; 13: sulfocysteine; 14: ethanolamine; 15: glycine and glycine-IS; 16: aspartic acid and aspartic acid-IS; **17:** citrulline; **18:** glutamic acid and glutamic acid-IS; **19:** sarcosine; 20: β-alanine; **21:** threonine and threonine-IS; **22:** homocitrulline; **23:** alanine and alanine-IS; **24:** γ-aminobutyric acid; **25:** hydroxylysine; **26:** aminoadipic acid; **27:** proline and proline-IS; **28:** glycil proline; **29:** β-aminobutyric acid; **30:** ornithine; **31:** cystathionine; **32:** α-aminobutyric acid; **33:** cystine and cystine-IS; **34:** lysine and lysine-IS; **35:** tyrosine and tyrosine-IS; **36:** methionine and methionine-IS; **37:** valine and valine-IS; **38:** homocystine; **39:** kynurenine; **40:** allo-isoleucine; **41:** isoleucine and isoleucine-IS; **42:** leucine and leucine-IS; **43:** phenylalanine and phenylalanine-IS; **44:** tryptophan and tryptophan-IS.

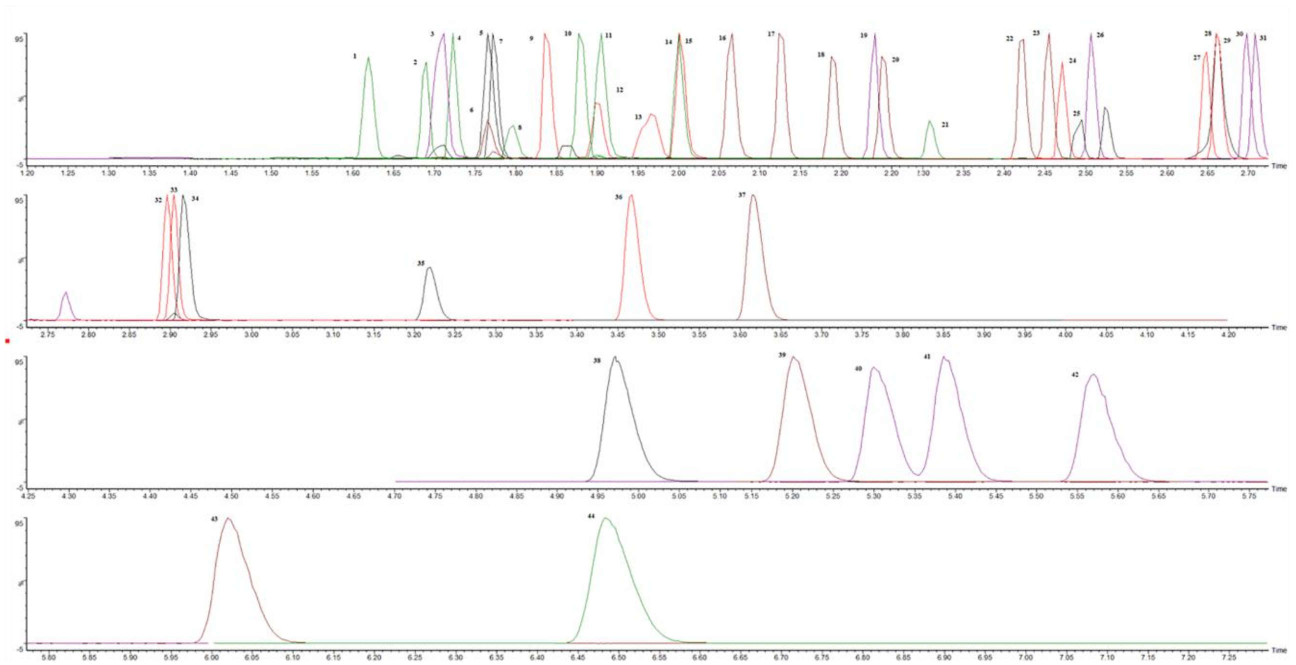

**Supplementary Figure 6.** Venn diagrams showing the representative metabolites examined using untargeted NMR metabolomics in conjunction with a targeted UPLC/MS approach. The two employed methods have 16 amino acid metabolites in common.

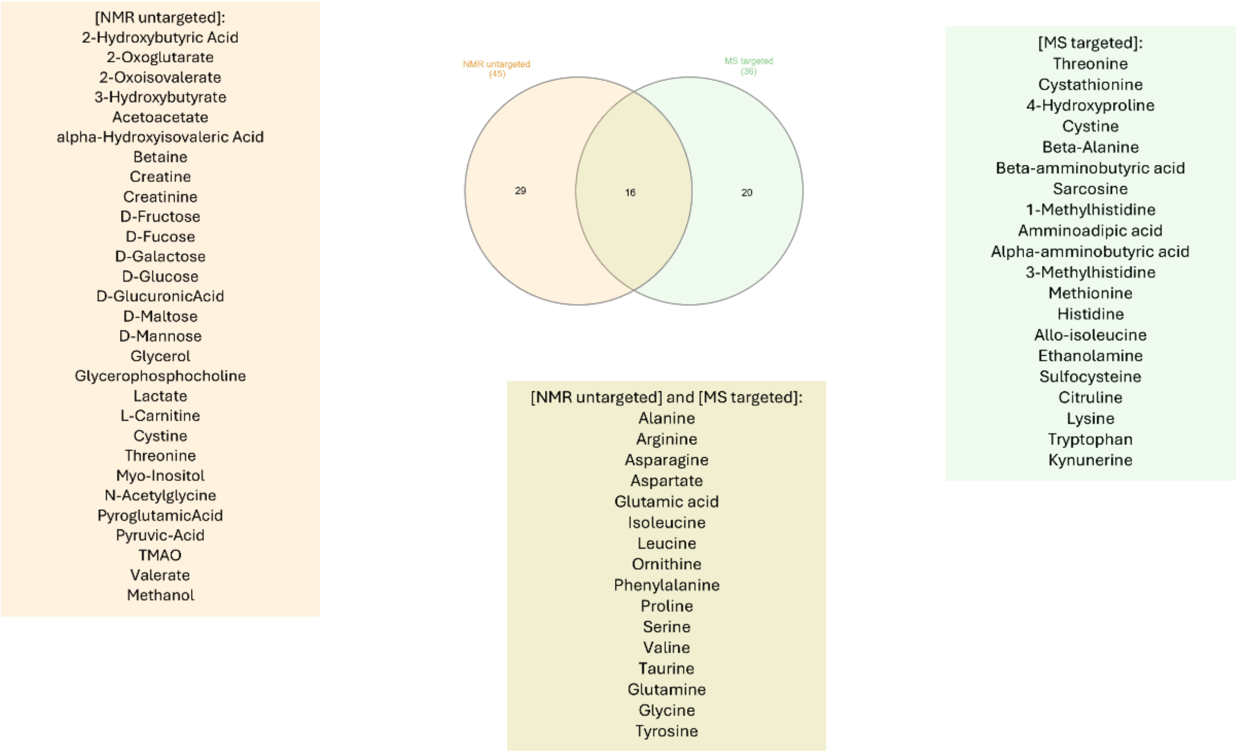


**Supplementary Figure 7. a**. Performance evaluations of the PLS-DA model related to UPLC/MS data by ROC curve. The ROC curve values for the first and second components with the relative p-value is given in the table **b**. Balanced error rate (BER) and total error (OVERALL) calculated on the first and second components using maximum, centroid and Mahalanobis distance.

**a**

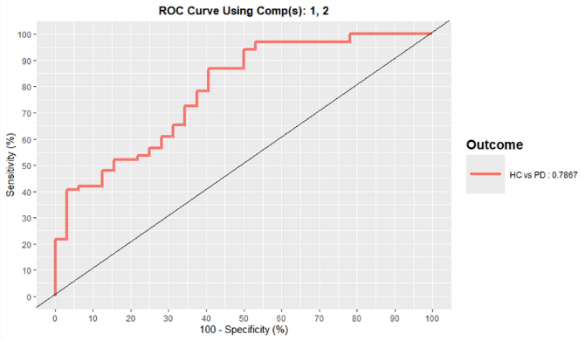


| AUC Component 1 | p-value |
|---|---|
| 0.73 | 0.00002 |
| **AUC Component 2** | **p-value** |
| 0.79 | 3.83e-6 |

**b**

| BER | MAX.DIST | CENTROID.DIST | MAHALANOBIS.DIST |
|---|---|---|---|
| COMP1 | 0,46 | 0.43 | 0,43 |
| COMP2 | 0.42 | 0,42 | 0,41 |
| **OVERALL** | **MAX.DIST** | **CENTROID.DIST** | **MAHALANOBIS.DIST** |
| COMP1 | 0,34 | 0,40 | 0,40 |
| COMP2 | 0,33 | 0,41 | 0,39 |

**Supplementary Figure 8.** Enrichment pathways analysis based on UPLC/MS data; the discriminative pathways between PD and HC are ranked according to p-value and number of hits reported in the bars. The hits represent the matched metabolites from the user-uploaded data; p-value describes the pathways' statistical significance index. In particular, Tryptophan metabolism (pvalue 1.12 e-2), Porphyrin metabolism (pvalue 3.33 e-2), and Glutathione metabolism (3.43 e-2).

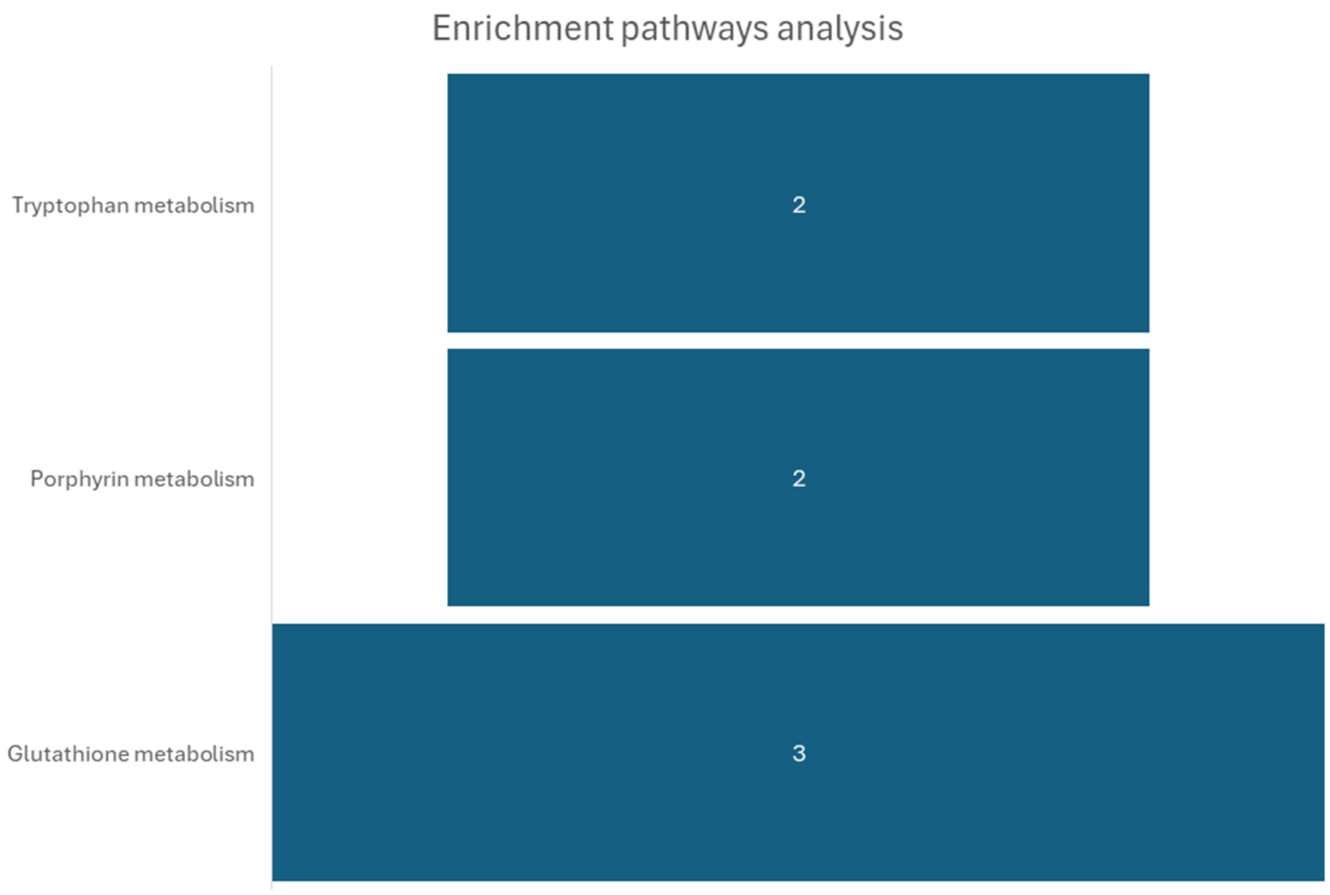

**Supplementary Figure 9. a.** The Correlation Matrix shows correlations between VIPs and clinical measurements using Pearson's correlation coefficient (r). Correlation values range from -1 (strong negative correlation, blue cells) to 1 (strong positive correlation, red cells). The data have been previous subjected to average normalization. **b.** Scatterplot showing the positive correlation between serum glycine concentration and MDS-UPDRS-III score, with an r value of 0.404 and the corresponding p-value 5.7 x$10^{-4}$ for statistical significance. Abbreviations: LEDD, Levodopa Equivalent Daily Dose; Mini N.A., Mini Nutritional Assessment; MMSE, Mini Mental State Examination; UPDRS-III, Unified Parkinson's Disease Rating Scale, part III.

**a**

| | Arginine | Aspartic acid | Cystathionine | Glycine | Glutamic acid | 4-hydroxypr. | Lysine | Ornithine | Threonine | Tryptophan | Valine | Kynurenine | Age | Mini N.A. | MMSE | Duration | LEDD | UPDRS III |
|---|---|---|---|---|---|---|---|---|---|---|---|---|---|---|---|---|---|---|
| Arginine | 1.000 | 0.062 | 0.228 | -0.048 | -0.230 | 0.060 | 0.253 | 0.383 | 0.279 | 0.217 | -0.140 | 0.076 | 0.001 | 0.050 | 0.165 | 0.004 | -0.039 | -0.015 |
| Aspartic acid | 0.062 | 1.000 | 0.092 | 0.067 | 0.108 | -0.115 | -0.251 | -0.248 | -0.226 | -0.060 | -0.214 | -0.074 | -0.181 | -0.053 | 0.131 | 0.299 | -0.112 | -0.145 |
| Cystathionine | 0.228 | 0.092 | 1.000 | 0.022 | -0.095 | 0.025 | 0.016 | 0.270 | 0.036 | -0.385 | -0.203 | 0.251 | 0.125 | -0.080 | 0.019 | -0.014 | 0.115 | 0.211 |
| Glycine | -0.048 | 0.067 | 0.022 | 1.000 | -0.008 | 0.196 | 0.054 | 0.075 | 0.236 | -0.162 | -0.224 | 0.174 | -0.042 | -0.171 | -0.122 | -0.003 | 0.256 | 0.404 |
| Glutamic acid | -0.230 | 0.108 | -0.095 | -0.008 | 1.000 | 0.072 | 0.110 | -0.076 | -0.035 | 0.160 | 0.446 | 0.043 | -0.096 | 0.077 | -0.083 | -0.015 | -0.144 | -0.126 |
| 4-hydroxypr. | 0.060 | -0.115 | 0.025 | 0.196 | 0.072 | 1.000 | 0.154 | 0.267 | -0.025 | 0.005 | 0.075 | 0.153 | 0.121 | -0.052 | -0.053 | -0.047 | 0.005 | -0.044 |
| Lysine | 0.253 | -0.251 | 0.016 | 0.054 | 0.110 | 0.154 | 1.000 | 0.601 | 0.552 | 0.516 | 0.573 | 0.199 | 0.076 | 0.152 | 0.105 | 0.183 | 0.125 | 0.042 |
| Ornithine | 0.383 | -0.248 | 0.270 | 0.075 | -0.076 | 0.267 | 0.601 | 1.000 | 0.528 | 0.296 | 0.290 | 0.285 | 0.012 | -0.012 | 0.005 | 0.142 | 0.166 | -0.036 |
| Threonine | 0.279 | -0.226 | 0.036 | 0.236 | -0.035 | -0.025 | 0.552 | 0.528 | 1.000 | 0.518 | 0.293 | 0.188 | -0.167 | 0.114 | 0.058 | 0.068 | 0.213 | 0.212 |
| Tryptophan | 0.217 | -0.060 | -0.385 | -0.162 | 0.160 | 0.005 | 0.516 | 0.296 | 0.518 | 1.000 | 0.583 | 0.049 | -0.233 | 0.235 | 0.183 | 0.055 | 0.071 | -0.200 |
| Valine | -0.140 | -0.214 | -0.203 | -0.224 | 0.446 | 0.075 | 0.573 | 0.290 | 0.293 | 0.583 | 1.000 | 0.023 | -0.035 | 0.234 | 0.053 | -0.027 | 0.063 | -0.140 |
| Kynurenine | 0.076 | -0.074 | 0.251 | 0.174 | 0.043 | 0.153 | 0.199 | 0.285 | 0.188 | 0.049 | 0.023 | 1.000 | 0.203 | -0.279 | 0.144 | 0.018 | 0.171 | 0.048 |
| Age | 0.001 | -0.181 | 0.125 | -0.042 | -0.096 | 0.121 | 0.076 | 0.012 | -0.167 | -0.233 | -0.035 | 0.203 | 1.000 | -0.152 | -0.057 | -0.095 | 0.007 | 0.152 |
| Mini N.A. | 0.050 | -0.053 | -0.080 | -0.171 | 0.077 | -0.052 | 0.152 | -0.012 | 0.114 | 0.235 | 0.234 | -0.279 | -0.152 | 1.000 | 0.178 | 0.079 | -0.079 | -0.019 |
| MMSE | 0.165 | 0.131 | 0.019 | -0.122 | -0.083 | -0.053 | 0.105 | 0.005 | 0.058 | 0.183 | 0.053 | 0.144 | -0.057 | 0.178 | 1.000 | -0.042 | -0.071 | -0.165 |
| Duration | 0.004 | 0.299 | -0.014 | -0.003 | -0.015 | -0.047 | 0.183 | 0.142 | 0.068 | 0.055 | -0.027 | 0.018 | -0.095 | 0.079 | -0.042 | 1.000 | 0.216 | -0.025 |
| LEDD | -0.039 | -0.112 | 0.115 | 0.256 | -0.144 | 0.005 | 0.125 | 0.166 | 0.213 | 0.071 | 0.063 | 0.171 | 0.007 | -0.079 | -0.071 | 0.216 | 1.000 | 0.263 |
| UPDRS III | -0.015 | -0.145 | 0.211 | 0.404 | -0.126 | -0.044 | 0.042 | -0.036 | 0.212 | -0.200 | -0.140 | 0.048 | 0.152 | -0.019 | -0.165 | -0.025 | 0.263 | 1.000 |

**b**

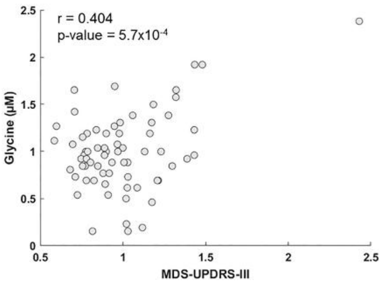
